# Comparison of predication of Acute Kidney Injury after cardiac surgery by health care providers versus the biomarker [TIMP-2]*[IGFBP7] and scoring systems. Protocol for a prospective single center observational study (PREDICTAKI)

**DOI:** 10.1101/2020.07.13.20148643

**Authors:** Wim Vandenberghe, Lien Van Laethem, Alexander Zarbock, Melanie Meersch, Eric A.J. Hoste

**Author notes:** Corresponding author: WimVandenberghe MD, ICU Ghent University Hospital, Ghent University.

## Abstract

**Introduction:** Acute kidney injury occurs in up to one third of patients after cardiac surgery and is an important contributor for adverse outcome. Previous research has demonstrated the benefit of a bundle of preventive measurements to reduce AKI in a subgroup of patients with high risk for AKI development. Urinary stress biomarkers [TIMP-2]*[IGFBP7] are used to identify these patients who are at risk for AKI. The trial aims to investigate the potential discrepancy between biomarker results and clinical estimation of occurrence of AKI on ICU in clinical practice.

**Methods and analysis:** We plan to include 100 adult patients after cardiac surgery with cardiopulmonary bypass in a prospective, single center clinical trial. After cardiac surgery, different type of healthcare professional in ICU will provide a prediction of AKI occurrence and severity in the next 48 hours by filling in a questionnaire just before and after [TIMP-2]*[IGFBP7] biomarker analysis. Primary, this trial investigates the potential discrepancy in AKI prediction between clinical estimation by healthcare providers, biomarker results, and previous described score systems. Secondly, the impact of knowledge of the biomarker result on the quality of prediction by healthcare providers will be evaluated.

**Ethics and dissemination:** This prospective, single center study has been approved by the medical ethical committee of the Ghent University Hospital (28^th^ May 2019, trial registration number B670201939991). Informed consent was obtained for patients and healthcare providers.

**Summary strength and limitations:**

- Influence of knowledge of a kidney biomarker on healthcare providers’ assessment of risk for AKI in clinical setting
- Different types of healthcare providers with various expertise
- It is a single center study with limited number of patients

## Introduction

Acute kidney injury (AKI) occurs in up to one third of patients after cardiac surgery, and is an important contributor for morbidity, mortality, and Health care utilization ^1^. In 2012, The Kidney Disease: Improving Global Outcomes (KDIGO) guideline provided a set of measurements to prevent AKI in high risk patients, the so-called “AKI Bundle” ^2^. Subsequently, Meersch et al showed in a single centre randomized trial a significant decrease in occurrence of AKI in a cohort of cardiac surgery patients when such an AKI bundle was used ^3^. To evaluate the effect of implementing the bundle in a multinational setting, a multicentre randomised controlled trial was performed by Zarbock et al (PrevAKI2) ^4^. In the PrevAKI trials, high risk patients for AKI were identified by the Nephrocheck® Test using a point-of-care device (Astute 140 meter) that provides results within 20 minutes (Astute Medical®). This test measures the two urine stress biomarkers tissue inhibitor of metalloproteinases-2 and insulin-like growth factor-binding protein 7 and reports these as the product of these biomarkers ([TIMP-2]*[IGFBP7]) ^5^. A cut-off of 0.3 is used to identify patients at high risk for AKI, and a value >2 identifies patients at highest risk for AKI ^6^.

In cardiac surgery patients, healthcare providers typically assess the risk for AKI by integrating information from diverse sources such as the information on previous health status of the patient, detailed information on the type of surgery, and the clinical course during surgery and the first few hours after ICU admission. This clinical AKI risk assessment is also influenced by experience and clinical expertise. There are also several scoring systems for prediction of occurrence of AKI after cardiac surgery. It is unclear whether the information on kidney stress provided by the NephroCheck® is complimentary to the assessment based on clinical expertise. We therefore want to investigate in this PREDICTAKI study the clinical risk assessment for AKI and compare this to the NephroCheck® AKI risk and scoring systems. In addition, we will evaluate the predictive performance of scoring systems for assessment of risk for AKI.

## Objectives and Aims

Aim 1: To investigate if healthcare providers are able to predict the occurrence of AKI and renal replacement therapy (RRT) 4-h after ICU admission

Null hypothesis: Healthcare providers are equal in predicting the occurrence of AKI and RRT within 48 hours after cardiac surgery compared to the biomarker [TIMP-2]*[IGFBP7] and scoring systems.

AIM 2: To investigate the impact of knowledge of the biomarker [TIMP-2]*[IGFBP7] result on the prediction of healthcare providers

Hypothesis 2: The prediction of healthcare providers will improve with the knowledge of the biomarker results.

AIM 3: Investigate if subgroups of healthcare providers are better in prediction of AKI and RRT at 48-h after cardiac surgery

Hypothesis 3: Physicians are better in prediction of AKI and RRT at 48-h after surgery in comparison to ICU nurses

## Methods and analysis

### Design and setting

This is a prospective, observational, single centre trial performed at the Ghent University Hospital.

### Patient and public involvement

Patients and public were not involved in the research design of the study. Study results will be published as an open access publication.

### Participants

Patients are eligible for inclusion if they are scheduled for cardiac surgery with the use of cardiopulmonary bypass, age is between 18 and 90 years and when written consent is obtained. Patients will be excluded if there is pre-existing AKI (≥ stage 1), need for cardiac assist devices (such as intra-aortic balloon pump (IABP), extracorporeal membrane oxygenation (ECMO), left ventricular assist device (LVAD), right ventricular assist device (RVAD)), when patients are pregnant or breast feeding, when there is known nephritis, interstitial nephritis or vasculitis, when there is chronic kidney disease (CKD) with eGFR below 20ml/min/1.73 m^2^, when there is need for RRT, or when the patient received a kidney transplant in the preceding 12 months.

### Co-enrolment in PrevAKI trial

The PrevAKI trial investigates the implementation of a bundle of supportive measures to prevent AKI in high risk patients in a multicentre setting. (Principal investigator: Prof. Dr. A. Zarbock, University Hospital Muenster, Germany, Belgium ethical committee project number: B670201734684). Co-enrolment in both PrevAKI and PREDICTAKI trials is allowed and the biomarker results will be used in both trials.

### Consent process

Written consent for the PREDICTAKI trial will be obtained according to the requirements of the local ethical committee. Consent will be asked to both patients and healthcare providers participating in the trial. Information will be given about the purpose of the trial, the design, the aim, its benefits as well as potential risks. Because this trial only involves one urine sample taken from the urinary catheter which is routinely placed for cardiac surgery, study related risks are absent. The consent will only be taken if the patient is able to give written consent, the day before surgery. All data will be kept and stored in pseudo-anonymised form. The patient has the right to withdraw from the study at any point, without reason.

### Trial workflow

Patients eligible for participation are asked for written consent after being thorough informed about trial purpose, design, aim, benefits and risks. Consent is asked one day before surgery.

Four hours after cardiopulmonary bypass for cardiac surgery, one urine sample is collected from the urinary catheter. [TIMP-2]*[IGFBP7] will be measured using the Nephrocheck® Test (Astute Medical). During Nephrocheck analysis, which takes about 20 minutes, questionnaires are filled in by several types of healthcare providers (HCP): 1. an intensivist, 2. a medical doctor in training on ICU, 3. an ICU nurse, and 4. a nephrologist (see figure 1 and 2). The first three HCPs are taking care of the patients, whereas the nephrologist is consulted almost simultaneously for risk assessment. Six AKI or renal replacement therapy (RRT) prediction scores are calculated: Cleveland Clinical score, Crate score, Mehta score, Murphy score, Ng Score and the ‘AKI predictor’ score ^7-12^. The results of these scores are not available for the HCPs when they fill out the questionnaire.

**Figure 1:**
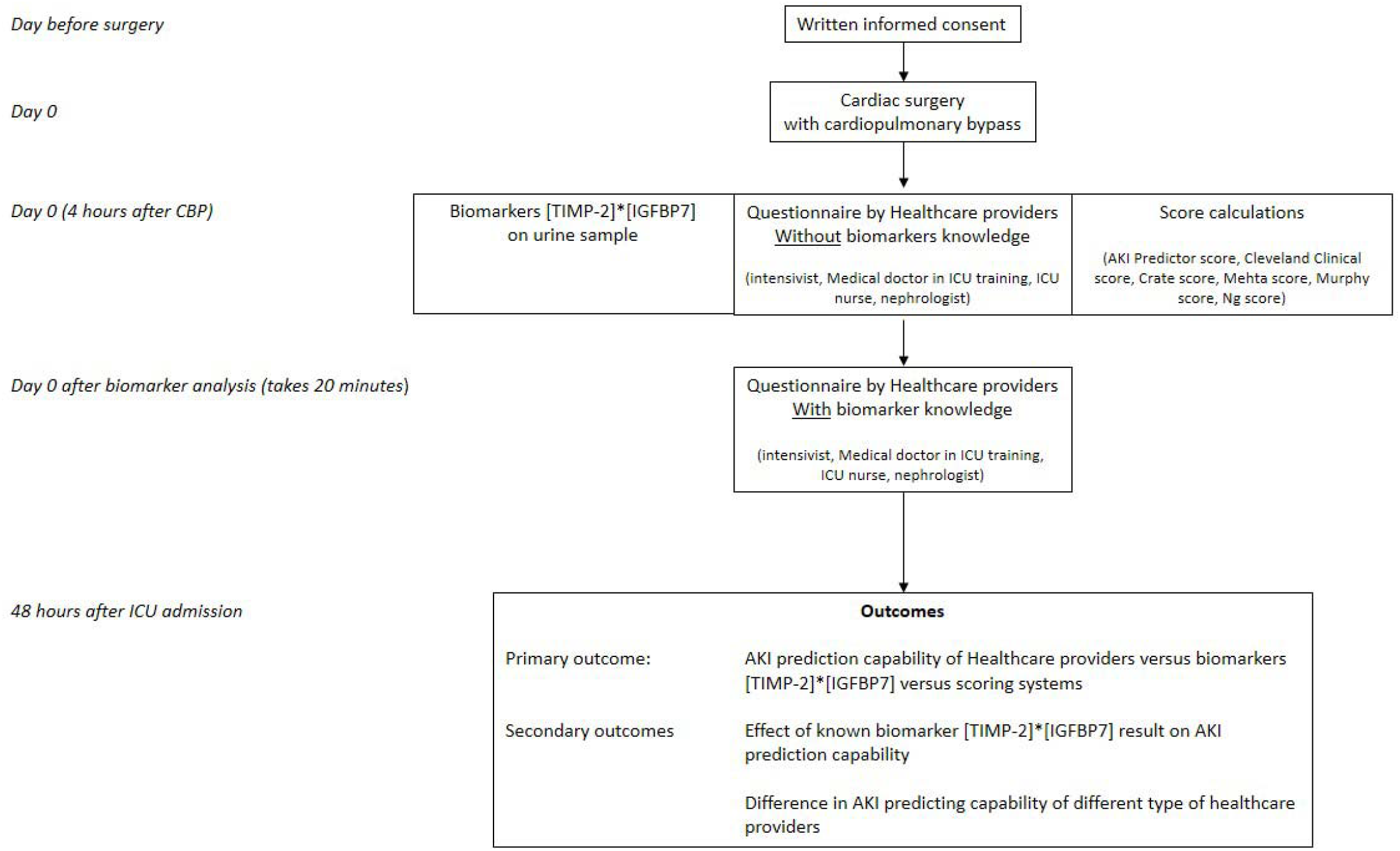
Trial workflow. Legend: AKI; acute kidney injury;Tissue inhibitor of metalloproteinases-2 and insulin-like growth factor-binding protein 7 ([TIMP-2]*[IGFBP7]); ICU, intensive care unit.

**Figure 2:**
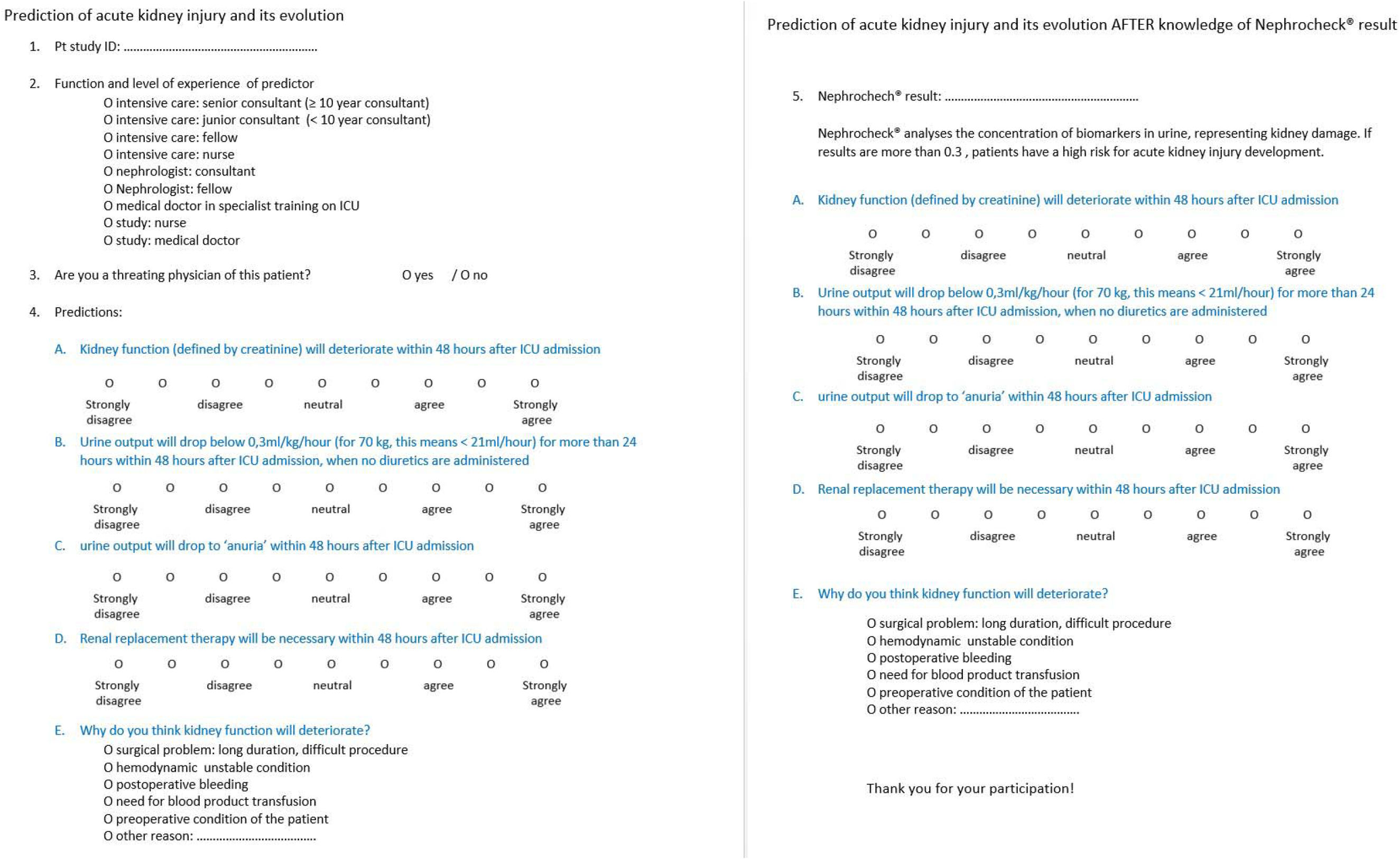
Questionnaire.

The questionnaire consists of 4 questions and answers are according to a 9 elements Likert scale (figure 2).

When the Nephrocheck® Test result is known, the result is presented to the HCPs and they are asked to again predict AKI, with the knowledge about the biomarker result.

All HCPs receive a short explanation (written and oral) on the Nephrocheck® Test result: a value ≥ 0.3 (ng/ml)^2^/1000 identifies patients with a high risk for AKI.

Kidney function will be followed up for 48 hours after ICU admission (See figure 1 Trial workflow). Demographic patient data will be collected.

### outcomes

#### Primary outcome

- Prediction of AKI and RRT 48 hours after ICU admission by HCPs, the NephroCheck® and 6 AKI scoring systems.

#### Secondary outcomes

- the influence of knowledge of the NephroCheck® test result, on the assessment of AKI risk by HCPs.
- the difference of AKI prediction by of several types of HCPs.

### Sample size

Since there are to the best of our knowledge no comparable studies on this topic, we could not perform a power calculation. We also acknowledge that clinical assessment of risk for AKI post cardiac surgery will be highly influenced by HCPs expertise and will therefore possibly vary in different settings. This pilot study is therefore designed to gain more knowledge on this specific topic, so that these data can be used to design future prospective multicentre studies. We arbitrarily set the number of patient inclusions for this trial at 100. For each patient, 4 HCPs will fill in a questionnaire before and after biomarker knowledge resulting in 800 questionnaires. Each questionnaire contains 4 predictions, resulting in a total of 3200 AKI related predictions.

### Statistical analysis

HCPs will fill in a paper questionnaire. These questionnaires and study specific data gathered by our electronic ICU patient data management system (PDMS) (Centricity Critical Care®, General Electric, USA) will be prospectively collected by using REDCap electronic data capture tools hosted at Ghent University.

Predictive ability will be evaluated with the receiver operator characteristic curve (ROC) and quantified by the area under the curve (AUC). AUC-ROC is defined as excellent when AUC-ROC ≥0.900 ; good: 0.800 ≤ AUC-ROC ≤0.899; fair: 0.700 ≤ AUC-ROC ≤0.799; poor: 0.600 ≤ AUC-ROC ≤ 0.699; and failed when AUC-ROC < 0.60. Calibration of the predictions will be evaluated. Interrater reliability will be investigated by calculation of percentage agreement, interclass correlation coefficient and Kappa statistics.

Quantitative parameters will be expressed as median and interquartile range (25^th^ to 75^th^ percentile) and compared using the Mann-Whitney U and Kruskal-Wallis tests. Qualitative parameters will be reported as number and percentage and compared using the Chi-square test. Two-sided P values of <0.05 will be considered statistically significant.

### Ethics and dissemination

This prospective, single center study has been approved by the medical ethical committee of Ghent University Hospital (28^th^ May 2019, B670201939991). Informed consent was obtained for patients and healthcare providers. Trial oversight will be performed by the Health, Innovation and Research institute of Ghent University Hospital (HIRUZ).

## Data Availability

Data is not public available, but can be provided on reasonable request

## Funding

There was no financial support for this study.

When patients are co-enrolled in the PrevAKI trial, Nephrocheck® device and cartridges were provided by this study.

## Competing interest

**WV**: non declared

**LV**: non declared

**AZ**: AZ received grants from Astute Medical, Baxter, Fresenius, and Astellas. He also received fees from Astute Medical, BioMerieux, Baxter, Fresenius, La Jolla Pharmaceuticals, and Astellas

**MM**: Astute Medical, Baxter, FMC: lecture fees

**EH**: AM Pharma: travel fee, Sopachem: speakers fee, Astute Medical: speakers fee, Alexion: speakers fee

## Authors contribution

**WV**: designed the trial, writing of the protocol, patients inclusions, design of Redcap Database, wrote the first draft

**LV**: designed the trial, patients’ inclusions, reviewing and revising of the manuscript

**AZ**: involved in trial design, reviewing of manuscript draft

**MM:** involved in trial design, reviewing of manuscript draft

**EH**: designed the trial, reviewing of manuscript draft

### Trial status

Recruitment started in 1^st^ July 2019 and estimate to complete the study in August 2020.

